# Impact of the COVID-19 pandemic on Latino families with Alzheimer’s disease and related dementias: Perceptions of family caregivers and primary care providers

**DOI:** 10.1101/2022.05.25.22275517

**Authors:** Jaime Perales-Puchalt, Jill Peltzer, Monica Fracachan-Cabrera, Adriana Perez, Mariana Ramirez-Mantilla, K Allen Greiner, Jeffrey M Burns

**Author notes:** **Corresponding Author:** Jaime Perales Puchalt, PhD, MPH, KU Alzheimer’s Disease Research Center, 4350 Shawnee Mission Parkway, Fairway, KS 66205. **Author contributions:** Jaime Perales-Puchalt made substantial contributions to the conception and design of the study, the acquisition, analysis, and interpretation of the data, and drafting and critically revising the manuscript for important intellectual content. Jill Peltzer made substantial contributions to the acquisition, analysis, and interpretation of the data and drafting and critically revising the manuscript for important intellectual content. Monica Fracachan-Cabrera made substantial contributions to the analysis, and interpretation of the data, and critically revising the manuscript for important intellectual content. Adriana Perez made substantial contributions to the acquisition and interpretation of the data and drafting and critically revising the manuscript for important intellectual content. Mariana Ramirez-Mantilla made substantial contributions to interpretation of the data and drafting and critically revising the manuscript for important intellectual content. K Allen Greiner made substantial contributions to the conception or design of the study, the interpretation of the data and drafting and critically revising the manuscript for important intellectual content. Jeffrey M Burns made substantial contributions to the conception or design of the study, the acquisition and interpretation of the data and drafting and critically revising the manuscript for important intellectual content.

## Abstract

Latinos experience disproportionately poor outcomes in dementia and COVID-19, which may synergistically impact their health. We explored the impact of the COVID-19 pandemic among Latino families with dementia via a qualitative descriptive study of 21 informal caregivers of Latinos with dementia and 24 primary care providers. Two themes arose: *The impact of a global pandemic* (e.g., accelerated cognitive and physical decline, or caregivers choosing between risking finances and the family’s infection given the work situation) and *Developing resilience to the effects of the pandemic (e.g., caregivers seeking vaccination sites, moving in with the care recipient and adopting telehealth)*.

## Introduction

Alzheimer’s disease and related dementias (ADRD) have a devastating impact on Latino families. Compared to non-Latino White individuals, Latinos experience disparities in ADRD risk, detection, treatment, and care. While Latinos are 1.5 times more likely to have ADRD,^1^ they are 1.4 times less likely to be diagnosed.^2^ Latinos are diagnosed with an eight-month delay, ^3^ are under-prescribed cholinesterase inhibitors,^4^ and underuse ADRD support services.^5^ These disparities are further compounded by Latinos’ lower ADRD knowledge and inclusion in ADRD research. ^6-8^

Latino families have also been disproportionately impacted by the COVID-19 pandemic. As of January of 2022, Latinos represent 8% of the US 65 and older population, but 13% of COVID-19 cases in the same age group.^9^ Latinos of any age are also more likely to be infected with COVID-19, representing 18% of the US population, but 25% of those infected. When controlling for age, the percentage of COVID-19 deaths is also higher among Latinos. For example, Latinos represent 14% of the deaths among those 65 and older, which is nearly twice as much as their representation in the US 65 and older population.^9^ Latinos are also less likely to be vaccinated against COVID-19, representing only 12% of those initiating COVID-19 vaccination.^10^ Some factors contributing to these disparities include having a job considered essential, living in a segregated geographic area, living in overcrowded households, limited English proficiency, and reduced use of preventive behaviors to avoid COVID-19 infection.^11^

Despite the growing literature on the impact of COVID-19 among Latinos, little is known about the impact of COVID-19 on Latino families with ADRD. Studying COVID-19’s impact on Latino families with ADRD is important given that Latinos are disparity populations in both ADRD and COVID-19, both of which might synergistically impact their health. ^9,10,12^ A study in California that included eight Latinos with cognitive impairment found that the biggest impact of COVID on these and other underserved communities included the fear generated by the pandemic, distress stemming from feeling extremely isolated, and receiving inaccurate information about COVID-19 from different sources.^13^ The study also reported some strategies participants used for coping during the pandemic (e.g., mask-wearing, remote communication), and the importance of access to essential resources such as friends, the church and local programs. While this work provided important insights on the impact and resilience factors for diverse populations with cognitive impairment, it is crucial to increase the Latino representation for a more detailed understanding. It is also important to listen to the perspectives of family caregivers and primary care providers (PCPs), which may provide a different point of view, allow triangulation, and represent individuals with more severe cognitive impairment. The aim of this study was to gain an in-depth understanding of the impact and resilience factors related to the COVID-19 pandemic among Latino families with ADRD. To achieve this goal, we interviewed family caregivers of Latinos with ADRD and PCPs who serve Latinos with ADRD across the US. Findings can inform actions to help Latino families with ADRD remain safe while maintaining a good quality of life during this ongoing pandemic and future ones.

## Materials and methods

This study used a qualitative descriptive design. Qualitative studies rely on text data and aim to understand the meaning of human action.^14^ The goal of this study was to gain an in-depth understanding of the impact of the COVID-19 pandemic on Latino families with ADRD. However, this study is part of a broader study that planned to identify what ADRD care services are offered in primary care and how they are delivered across a variety of settings.

The University of Kansas Medical Center Institutional Review Board approved this project (STUDY00145615).

We recruited two groups of participants: 1) family caregivers of people living with ADRD and 2) PCPs. Inclusion criteria for caregivers included being 18 or older, identifying as a close friend or relative of a Latino person with ADRD diagnosed by a healthcare provider or research study, providing, or having recently provided care to them at least once a week in-person or via phone, being proficient in English or Spanish, living in the US, and being willing to participate in the study. Inclusion criteria for PCPs included being a medical doctor, doctor in osteopathic medicine, nurse practitioner, or physician assistant, who currently or recently provided primary care services to Latino families with ADRD in the US.

Caregivers were recruited via convenience sampling from diverse sources, including a research registry, clinic, and community ADRD patient lists, internet and newspaper advertisements, and connections with community partners and research team members. PCPs were initially recruited using snowball sampling techniques, first contacting connections of the research team and later asking interviewed PCPs for referral of other PCPs.

We used purely qualitative semi-structured interviews, which allow for the comparison between participants, while allowing spontaneous exploration of topics relevant to unique participants.^15^ The interviews took place between November 2020 and April 2021. All interviews but two were conducted via secure videoconference. The other two interviews were conducted over the phone. Before the interview, all participants completed an informed written consent online either via their computers, tablets, or phones.

The process for each interview was similar: the first author interviewed all participants. The interview started with a short conversation aimed at developing rapport and explain the main goals of the interview. The first questions asked about basic characteristics of the participants. Core questions of the survey asked about participants’ experience with primary care clinics (caregivers) and serving Latinos with ADRD (PCPs). Unless participants had mentioned it spontaneously, the interviewer asked participants halfway through the interview how the COVID-19 pandemic had impacted them and their care recipient (caregivers) and the Latinos families with ADRD they serve (PCPs). The interviewer audiotaped all interviews, which were designed to last 45-60 minutes. Interviews were in English or Spanish and those with caregivers included simple language to account for different levels of literacy. A professional team transcribed all interviews and the interviewer reviewed them for accuracy. The research team compensated participant’s time by posting them a $40 gift card.

We organized the transcripts for qualitative review, using a pragmatic approach and thematic analysis methods.^16-18^ We organized the data in DeDoose.^19^ Both authors read the interviews a first time to familiarize themselves with the data. The first author identified relevant sociodemographic, ADRD relationship, and clinical service data in the interviews and summarized it into descriptive statistics. The fist author also condensed the meaningful bits of text into shorter text and developed codes (1-3 descriptive words) using an iterative process reading the transcripts line by line. After condensing and coding the transcripts, the first author developed subthemes within the text by iteratively contrasting codes and later developed themes by contrasting subthemes. Two researchers (JPP and MFC) conducted independent reviews of the codes, themes and subthemes and resolved coding disagreements through discussion and consensus. To make bring rigor and validity to the research process, the interviewer used active listening techniques during the interview aimed at confirming the information shared by participants. The interviewer also emphasized the fact that participants were the experts in their experiences to reduce power differentials. Both coders had previous coding experience. The fact that only one coder conducted the interviews and they both had different educational backgrounds allowed different perspectives.

## Results

### Characteristics of the sample

Table 1 shows the characteristics of the 21 family caregivers and 23 PCPs. Most caregivers were women (90.5%), younger than 66 (76.2%), and children (71.4%) of people with Alzheimer’s disease (66.7%). All but one participant identified as Latino of diverse origins, 28.6% were born in the US, and all lived in urban regions from the Midwest (71.4%), Northeast (23.8%) and the South (4.8%). Interviews were conducted in Spanish with 61.9% of caregivers, 66.7% of them reported good English proficiency, and 38.1% of them reported their care recipient had good English proficiency.

**Table 1.** Characteristics of the sample.

|  | Family caregivers (n=21) |  | Primary care providers (n=23) |  |
| --- | --- | --- | --- | --- |
|  | % | n | % | n |
| Women | 90.5 | 19 | 56.5 | 13 |
| Age |  |  |  |  |
| 33-65 | 76.2 | 16 | - | - |
| 66+ | 23.8 | 5 | - | - |
| Ethnoracial group |  |  |  |  |
| Latino | 95.2 | 20 | 43.5 | 10 |
| Non-Latino White | 4.8 | 1 | 43.5 | 10 |
| Non-Latino Black | 0.0 | 0 | 8.7 | 2 |
| Non-Latino Asian | 0.0 | 0 | 4.3 | 1 |
| Subgroup among Latino participants |  |  |  |  |
| Mexican | 50.0 | 10 | 20.0 | 2 |
| Caribbean | 15.0 | 3 | 30.0 | 3 |
| South American | 15.0 | 3 | 30.0 | 3 |
| Central American | 20.0 | 4 | 10.0 | 1 |
| US-born | 28.6 | 6 | 52.2 | 2 |
| Urban setting | 100.0 | 21 | 78.3 | 18 |
| Interview in Spanish | 61.9 | 13 | 21.7 | 5 |
| Region |  |  |  |  |
| Midwest | 71.4 | 15 | 65.2 | 15 |
| Northeast | 23.8 | 5 | 8.7 | 2 |
| West | 0.0 | 0 | 17.3 | 4 |
| South | 4.8 | 1 | 0.0 | 0 |
| Puerto Rico | 0.0 | 0 | 8.7 | 2 |
| Good English proficiency | 66.7 | 14 | - | - |
| Can provide services in Spanish | - | - | 52.2 | 12 |
| Relation to care recipient |  |  |  |  |
| Child | 71.4 | 15 | - | - |
| Spouse | 23.8 | 5 | - | - |
| Friend | 4.8 | 1 | - | - |
| Diagnosis of person with care recipient |  |  | - | - |
| Alzheimer's disease | 66.7 | 14 | - | - |
| Dementia (unspecified) | 9.5 | 2 | - | - |
| Mild cognitive impairment | 9.5 | 2 |  |  |
| Early onset Alzheimer's disease | 4.8 | 1 | - | - |
| Early onset Alzheimer's disease and Frontotemporal dementia | 4.8 | 1 | - | - |
| Parkinson's dementia | 4.8 | 1 | - | - |
| More than one care recipient | 4.8 | 1 |  |  |
| Good English proficiency of care | 38.1 | 8 | - | - |
| recipient |  |  |  |  |
| Type of provider |  |  |  |  |
| Medical doctor | - | - | 65.2 | 15 |
| Nurse practitioner | - | - | 30.4 | 7 |
| Physician assistant | - | - | 4.3 | 1 |
| Type of clinic |  |  |  |  |
| Private | - | - | 47.8 | 11 |
| Safety net and federally qualified | - | - | 34.9 | 8 |
| Academic | - | - | 17.3 | 4 |
| Percent of Latino patients by provider |  |  |  |  |
| Less than 29% | - | - | 30.4 | 7 |
| 30-74% | - | - | 30.4 | 7 |
| 75%+ | - | - | 39.1 | 9 |
| Minutes of recording: Total; Min-Max | 1079 | 25-61 | 1162 | 35-73 |

More than half of PCP participants were women (56.5%), US-born (52.2%), Latino (43.5%) of diverse origins and non-Latino White (43.5%). Most PCPs were from urban settings (78.3%), with the Midwest being the most common region (65.2%). Approximately half of PCPs (52.2%) reported being able to communicate with their patients in Spanish. Most PCPs were medical doctors (65.2%) and nurse practitioners (30.4%) who reported a variation in the types of clinics they practiced in with a wide distribution of Latino patients served.

### Themes

We developed two themes from the analysis: T*he impact of a global pandemic* and *Developing resilience to the effects of the pandemic*.

Table 2 shows the themes, subthemes, and codes identified in the qualitative analyses.

**Table 2.** Themes, subthemes, and related codes.

| Theme | Subtheme | Code |
| --- | --- | --- |
| The impact of a global pandemic |  |  |
|  | Physical |  |
|  |  | Infection |
|  |  | Accelerated deterioration |
|  |  | Poor nutrition |
|  |  | Death |
|  | Psychological |  |
|  |  | Fear of infection |
|  |  | Uncertainty |
|  |  | Stress |
|  |  | Depression |
|  | Social |  |
|  |  | Isolation |
|  |  | Engagement |
|  |  | Work |
|  |  | Family relationships |
|  | Other |  |
|  |  | Unspecific negative |
| Developing resilience to the effects of the pandemic |  |  |
|  | Minimizing risk for COVID-19 |  |
|  |  | For physical consequences |
|  |  | For psychosocial consequences |
|  |  | Barriers |
|  | Social support |  |
|  |  | Informal support |
|  |  | Formal support |
|  |  | Facilitators and barriers |
|  |  | Consequences |
|  | Remote communication |  |
|  |  | Use |
|  |  | Facilitators and barriers |
|  |  | Consequences |

Theme 1. T*he impact of a global pandemic* describes the fear, stress, exhaustion, and unanticipated challenges that impacted individuals with ADRD, their caregivers, and their providers. Care recipients and caregivers had to adapt their daily schedules to promote a safe environment for the care recipients, who were at increased risk for COVID-19 and its sequalae.

1) Physical impact. Caregivers and PCPs reported multiple ways physical health was threatened or declined. First, the fear of COVID-19 infection for the care recipient was reported by most of the caregivers and PCPs. While some caregivers mentioned they or their care recipients had not been infected, they were fearful about the risk of COVID-19 to the care recipient, who was at risk for significant disease sequalae. Other caregivers or their care recipient (n=5) had been infected or were indeed infected during the interview. PCPs also mentioned their patients with ADRD acquiring COVID-19. As an extreme case, one PCP who serves institutionalized patients reported:

> *“One of my nursing homes right now, I think we are up to 22 positives today out of 40 patients”*

Second, caregivers and PCPs noticed accelerated cognitive and physical deterioration. One of the caregivers had to move their loved one to a long-term care facility in the midst of the pandemic and saw a rapid decline in cognition. Another PCP reported how the lockdown resulted in reduced access to treatment among some patients leading to accelerated cognitive and physical decline. The caregivers and PCPs attributed decline to social isolation, and lack of engagement in activities that affected mood, such as depression, sadness, and apathy. A caregiver explained:

> *“[Explains how after the lockdown, the care recipient only remembers long term memories]. So, I was thinking that all the time she was locked down here because of the cold weather and COVID might have affected her more”*.

Caregivers’ and care recipients’ fear of infection reduced in-person primary care visits, and hindered depression treatment or ADRD assessment. PCPs had to reduce physical contact with care recipients, which reduced their chance to convey warmth to their patients.

The pandemic influenced availability and quality of services, such as educating Latino caregivers on ADRD, assessing for dehydration, creating rapport, and some caregivers reported confusion among their clinicians in the identification of COVID-19 vs ADRD symptoms:

> *“When [care recipient] got the virus, she could not swallow. She had huge difficulty. We called the PCP and the neurologist, and we got an appointment later that day. Both the PCP and the neurologist saw the issue as part of the ADRD, not as part of the virus. Both independently. They said -Your [care recipient] is in her last days… let her rest, don’t insist-But I’m sure it was the virus because she could swallow again after two or three weeks, and she was doing better than before”*.
>
> Another caregiver shared *“My [care recipient] got COVID-19 in May. In September, I started telling her PCP that she had become very forgetful. She told me those were side effects of COVID and that little by little her memory would come back. My [care recipient] has always had a great memory. She’d never forget anything. This time the change was more drastic and that’s why I kept insisting to her PCP that it was something else. Every three months, I’d go to her and insist, until a neurologist saw her and diagnosed her”*.

Third, food access was a critical issue leading to poor nutrition that contributed to accelerated declines in health. Reasons for food access were due to delays in the mailing system and financial insecurity. This affected not only the care recipients but the caregivers. For example, the relative of a care recipient explained:

> *“[Going to restaurants] is one thing she cannot do anymore, and my cooking abilities is not all that great…, so fast-food has been our life for the last year maybe even more because she has not been able to cook for some time”*.

Fourth, participants reported death as a consequence of the pandemic among their Latino families. For example, a PCP explained that several of his Latino patients died during their yearly visit to their home country; deaths that would not have occurred prior to the pandemic. A caregiver explained how a relative with ADRD died short after being moved to a residential facility.

> *“…he was living alone*… *He was still early stage and he started getting psychotic episodes…, so I lived with him for about a month to try to figure out the medications, but he was very violent, so we had to put him in the hospital… for about a month and then you know I was able to find a residence for him, but he did not live too long after that, he literally stopped eating*…, *so eventually you know he was put on Morphine because he was in a lot of pain from multiple things and that is how he passed*.*”*

2) Psychological impact. Caregivers and PCPs reported multiple ways the pandemic influence mental and emotional health. First, participants often mentioned their and their care recipients’ fear of the care recipient becoming infected with COVID-19. A PCP described this fear as follows:

> *“Most dementia patients are old and this is the population at the highest risk, and they are even more scared [of getting the virus]”*.

This fear was not unfounded. Prior to the availability of the vaccine, caregivers and care recipients acquired COVID-19. As Latino older adults, they were at an increased risk for complications including death, causing significant chronic concern and fear. A PCP said:

> *“They do not want to come in. The news said very early that ethnic minorities were getting more severe disease that they just stopped coming in and it is really hard to get a hold of them”*.

Second, according to both caregivers and PCPs, caregivers and care recipients had feelings of uncertainty. There was uncertainty about when they would be eligible to get the COVID-19 vaccine, when the caregiver would get some respite by being able to take the care recipient back to the senior center, confusion about what was happening due to cognitive impairment, and general uncertainty about their lives. One caregiver defined their lives during the pandemic succinctly as:

> *“Everything has been in limbo with COVID”*.

Third, heightened fear and daily uncertainty led to increased levels of recurring stress. On top of pandemic-related stress, caregivers were unable to access caregiver support services and the respite they needed to manage their own health. They also lacked access to care residences needed to provide the safest care for the care recipients. This stress adds on top of the other stresses Latino families often experience, as this caregiver explained:

> *“I was recently told I had [a serious disease]. Now I need to get some further testing. This affected me a lot too. It’s also been hard to work, take care of the house, my kids… because when it’s not one problem it’s another. It’s been really hard… so much stress”*.

Fourth, care recipients and PCPs highlighted the frequency and severity of depressed mood among caregivers and care recipients, especially during the lockdown due to lack of social support and social isolation. The PCPs noted that lack of social support and social isolation due to lockdown negatively impacted mood, sharing:

> *“Umm, all of my ADRD patients are sad*… *they are isolated, by themselves”. “Social isolation of COVID has caused a significant level of depression”*.
>
> A caregiver also shared, *“And at this point I’m very sad, depressed, neurotic… I’ve no patience… very little patience. To me, life has changed a huge deal… 360 degrees”*.

3) Social impact. Caregivers and PCPs reported multiple social impacts on the care recipients and caregivers. First, participants, while understanding the need for the lockdown, frequently described how the lockdown contributed to social isolation that reduced engagement in activities and led to depression and apathy. The pandemic paralyzed or slowed down operations in clinics, church services, caregiver support activities, senior centers, and residences. Additionally, PCPs, home health assistants, residence staff and family members had gotten infected. In fact, a caregiver’s former PCP who spoke Spanish had died of COVID and was replaced by a non-Spanish speaker.

One of the PCPs mentioned:

> *“I think it is really a negative impact and really socially isolating a lot of people, which as a PCP always makes me worried because of how much stress that adds to people”*

Second, caregivers also reported that their care recipient and themselves lost access to engaging activities during the confinement, such as visiting with their peers, going out to eat, and attending social events at the local senior center. A caregiver summarized this feeling as follows:

> *“We have been so stuck with this life of COVID not being able to do anything or go anywhere”*.

Third, the pandemic interrupted work among caregivers. While this interruption has given some the opportunity to care for their loved ones, the inability to work had significant impact on the family’s financial situation. For some participants, the work environment increased the risk for COVID-19, as they were considered essential workers and had to face clients.

Families had to make difficult choices about exposing frail older members to COVID-19 or lose critical income to support the family. A PCP explained how it is affecting some Latino caregivers negatively:

> *“The pandemic has really affected the population. Some people have not been able to work. Many of them work in restaurants and similar jobs that have been affected by it. In construction, people have had no issues, but they have a lot more work. I have a large patient population that work in the meat packing plan industry, and they were really scared of COVID infection due to an outbreak they experienced”*.

Fourth, the pandemic negatively impacted highly important family connections. Family visits were reduced in both quantity and quality critical to social support. In addition to limited family gatherings, conflicts were created or exacerbated due to family members’ divergent opinions about pandemic guidelines and own personal needs. For example, the family caregiver of a care recipient would argue with another relative due to different perspectives on the care recipient not wanting to wear the face mask. A caregiver who moved back from another country to live with her *[care recipient]* shared that another relative was angry she made the move, stating *“I ended up fighting with my relative, who was also in [another country] because I ended up moving with [care recipient]. I told her, who will care for [care recipient] … our [care recipient]?”*

Another family caregiver who limited her visits to reduce the risk of infecting her care recipient mentioned:

> *“Well, I think that it has been incredibly difficult for her, for me, and for everyone because it is sad to say for whatever reason… I try to keep the peace with my [other relatives], so I gave her an iPad, but she does not know how to use it and she needs help using it and he is reluctant to facilitate the facetime calls”*.

4) Other impacts. In addition to the biopsychosocial impacts of the pandemic, caregivers and PCPs also expressed other perceived or expected impacts on care recipients. Participants have used unspecific terms to define the pandemic such as being very difficult, impacting greatly, or getting worse. Some PCPs suspected they had only seen the superficial issues with the pandemic and manifested concern about potential long-term consequences. Two PCPs mentioned:

> *“It is going to be almost a year in the next couple of months, so we have yet to determine the full impact on our patients because our one-on-one interactions have been very limited. I think there are other consequences or other issues that we will have to deal with as a result of not being able to see them because every state had its way, everybody closed, and some managed their re-openings differently”*.
>
> *“I do not know that I have seen the fallout yet… I think as time comes and things continue to hopefully settle as vaccines become widespread, people start getting more and more comfortable with getting back to the office for what they would consider non-emergent or non-urgent care, then we’ll start to see things that have been shuffled under the rug for 12 months or how long it is, until we start seeing those things kind of come near their face”*.

Theme 2. *Developing resilience to the effects of the pandemic* describes the strategies that care recipients and caregivers used to adapt to the pandemic to manage their daily lives and their health. This theme also conveys how the PCPs navigated the pandemic to support families living with ADRD.

1) Minimizing risk for COVID-19. First, to mitigate physical consequences, multiple caregivers and care recipients acquired vaccinations as soon as they were available. Getting vaccinated also allowed the family to provide care for their loved ones rather than avoiding contact. PCPs were proactive in ensuring their patients were aware of the vaccines. One PCP shared that the Latino families were more receptive to vaccines than were the Anglo patients. He believed that the Latino families’ higher sense of susceptibility to and severity of COVID led to their early uptake of the vaccine and other COVID-19 prevention measures:

> *“There are very few Latinos that deny COVID-19. I feel like this happens more often among Anglos. Especially before the election, it was crazy. It was more about politics than anything. But my Latino patients, most understand there’s an issue because they know how it’s affecting people in their countries of origin”*.

Caregivers also made sure they, and their care recipients would wear face masks and shields, wash their hands often, kept a safe distance from others (or themselves if the caregiver was exposed to others), and got tested for COVID-19. Caregivers took over some tasks they typically did not do, to increase the care recipient’s safety and physical health, including taking care of medicines, nutrition, planning activities and going shopping alone. Some caregivers were particularly proactive, as they worried that their usual healthcare provider was taking too long to give care recipients their vaccine. Two caregivers shared:

> *“I feel like if I had not been persistent and kept calling, she probably would still not have her vaccines*.*”*
>
> *“So, we found some other sources and I do not understand how such an old person has not gotten a call from her primary place, you know… So, I asked [nun at a catholic hospital at a different state] if she could give permission for my [care recipient] to go and she did and so my [care recipient] got vaccinated in [that other state]”*.

Second, caregivers also found ways to mitigate the psychosocial consequences of the pandemic. One family who lived in different states got vaccinated to hold a family gathering. The participants found ways to adapt to being home for long periods of time. Participants shared that adapting to being at home has buffered against stress and allowed families to function better during the pandemic. Caregivers also shared strategies, like explaining the pandemic to the care recipient, requesting patience from formal caregivers by explaining the effect of the pandemic on the care recipient, and staying at home even when a senior center reopened. A caregiver described:

> *“The senior center opened two weeks ago. But I told them I’d have her at home. There’s no need to expose her to anything… Four hours are not going to make a difference and it’s ultimately going to give me more work. And she would be exposed. I don’t trust that the protocols are ideal for her to go*.*”*

Third, despite employing strategies to manage psychosocial consequences, participants continued to face challenges that included cognitive issues of the care recipient that hindered understanding, the resistance of the care recipient to wear the face mask, and issues with the operations of clinics including not having available vaccines or not returning calls. This spousal caregiver experienced the following:

> *“I mean they have been supportive in everything. One thing that upset me a little bit was I got an e-mail not too long ago from the hospital saying that if I wanted the COVID shot to call and make an appointment and when I did to make my appointment, I said what about my [care recipient] and they said her name has not come up yet and I said she needs it more than I do. I called the neurology clinic asking if they could intervene and help me, they never returned my call*.*”*

2) Social support was critical for reducing social isolation and its sequalae. First, caregivers sought informal sources of support, including family and friends, and formal sources, such as clinical resources and respite care. Family support was the most frequently mentioned type of social support. Many child caregivers either moved in with the care recipient or moved the care recipient with them to provide daily care. There were only few cases where a caregiver reported not getting family support to the care recipient:

> *“I am the only one that visited her and would take her anywhere. No one has visited ever since I moved back. My [other relative] might have visited once, maybe*.*”*

Other types of family support included the caregivers taking the care recipient to another relative’s home for a few hours for care and engagement, visiting or calling more often, or scheduling healthcare visits. Other relatives were also a source of emotional support, for example, kids and grandkids coming over to visit. Friend support included neighbors who would assist care recipients in things such as contacting a professional when they locked themselves out or caregiving support program members that continued to be in contact despite support groups having halted during the pandemic:

> *“The reason why I’m not going to support groups now is initially because of COVID. They had stopped and I had not heard that they are doing anything with the group again, but I have been in contact with several of the people that I met through the group”*.

Second, there were four sources of formal social support: respite care, clinics, senior centers, and homecare attendants. Caregivers most often sought help from their providers and clinical staff. Clinics provided support by ensuring clinicians and staff were vaccinated and enacting other precaution measures (e.g., masks, distance, hand washing), increasing curbside visits, organizing medication refills with pharmacies to avoid patients having to attend the clinics, encouraging families to stay at home, providing COVID-19 vaccines, doing extra visits to reduce care recipients’ sadness, and encouraging patients who live alone or are sad to use their counseling services. A PCP mentioned the importance of not waiting for patients to make the appointments, but use a rather proactive outreach strategy:

> *“COVID has been major issue now that people do not get to go to places, so our outreach is more important now because they are really missing, they are not just now isolated, now they also might be feeling lonely or not being able to see grand kids or family… and as a result I think that has really affected them*.*”*

Four care recipients lived in residences with different levels of assistance. Two moved during the pandemic and the other two already lived the before the pandemic started. Most senior centers were closed during the pandemic, but some all-inclusive care programs for the elderly reopened shortly after the lockdown was lifted. Several caregivers also relied on home care professionals to care for their care recipients. Caregivers were willing to pay for home care out of pocket and several also had created trust funds for the expensive care needs of their loved ones. Home care services mostly came from private sources, but also included all-inclusive care programs for the elderly and catholic charities. Regarding the all-inclusive care programs for the elderly, a caregiver and a PCP mentioned:

> *“The home care services come from [an* all-inclusive care program for the elderly]. *I think since [month] from last year they’ve been coming every day from [specific time] to [specific time], which is when I have more work online meetings”*.
>
> *“Now if they need to be seen we send people to the home and we can do that. If somebody needs to be seen we will send a provider, a nurse, whoever else needs to see them”*.

Third, there were several facilitators and barriers of support at the clinic level, including a shorter clinic distance to care recipients’ homes, clinic COVID-related precautions, cultural tailoring, and flexibility in the visit modality (in-person and remote as needed). PCPs also mentioned politics, public health professionals and scientists’ involvement in getting funding to develop and implement the COVID-19 vaccines.

Regarding cultural tailoring, a caregiver mentioned:

> *“You know we had meals on wheels, but we just could not have those meals, they were terrible. They were terrible for a Latino family”*.

Fourth, consequences of social support were physical, psychological, and social.

Examples of physical consequences include potentially reducing mortality by providing formal and informal caregiving services. Psychological consequences include clinic and family support reducing loneliness and increasing feelings of safety. Social consequences include caregivers being allowed to accompany their care recipients during clinic visits, curbside visits allowing socialization and home care services lowering isolation.

3) Remote communication. Remote communication is a potential facilitator to social support. However, given the high frequency of comments by caregivers and PCPs, we decided to create an independent category. First, participants stressed their increase in the use of remote communication during the pandemic. Purposes of remote communication included remote work, communicating with the care recipient and other relatives, conducting telehealth between clinicians, caregivers, and care recipients, and using interpreters. These communications included phone and video calls, as well as emailing and patient portals. This PCP exemplifies this shift:

> *“That’s the way I’ve communicated with patients during the pandemic… over the phone or virtually and ordering their medications directly electronically to the pharmacy or the lab”*.

Second, there were several barriers and facilitators to remote communication. Facilitators to using telehealth among patients include social engagement, and clinician flexibility with using different modalities according to the families’ needs and abilities (e.g., requesting the patient to talk with the caregiver or using speakerphone, phone call, video call or patient portal). Barriers include care recipients having cognitive or physical impairment, especially when there is no family involvement. The relative of a care recipient said:

> *“…but it is really hard to Facetime with him unless there is somebody right there with him because the camera keeps going to the ceiling or he does not want to talk to me, so having a Facetime with him is sometimes difficult”*.

A barrier to remote communication was language communication difficulties (e.g., patient portals not being in Spanish, translators being harder to understand over the phone vs in-person). A PCP also identified trust of the remote communication source, and low technology availability and savviness as barriers specifically among the Latino families they serve. This example includes both barriers:

“I have seen specifically in the Latino population they are not quite as engaged with some of the technology and telehealth, I think they are a little bit worried about how to use it or you know is my information safe”.

Third, consequences of the higher use of remote communication were both positive and negative. Positive consequences include a higher access to services (e.g., primary care, counseling, neurology, Alzheimer’s Association), higher access to PCP training, and care recipient engagement with activities. Telehealth was helpful in reducing levels of stress when it worked well. However, remote communication also had negative consequences. When remote communication did not work well, it was a source of stress. These are two examples of positive and negative consequences linked to stress and virtual communication reported by a PCP and the relative of a care recipient who does not live with her:

> *“The ADRD patients don’t do the greatest with video, I am learning. It’s very confusing and upsetting to them”*.
>
> *“When I am not involved [in calling care recipient often] she is just very anxious, so I think calling multiple times a day everyday helps”*.

## Discussion

This study aimed to gain an in-depth understanding of the impact and resilience factors related to the COVID-19 pandemic among Latino families with ADRD, from the perspectives of family caregivers and PCPs. To achieve this goal, we interviewed 21 family caregivers of Latinos with ADRD and 23 PCPs who serve Latinos with ADRD across the US. These participants were diverse with respect to their region, primary language, and other characteristics. We found that the COVID-19 pandemic has impacted Latino families with ADRD at physical, psychological, social, and other levels. To address that impact, Latino families with ADRD have used a range of resilience strategies.

COVID-19 has had an important impact on Latino families with ADRD at many levels. Physically, our participants’ comments about infection and death are consistent with the Latino COVID-19 disparities reported by the Centers for Disease Control.^9^ This impact was experienced disproportionately in nursing homes, which use is lower among Latinos,^20^ but where a higher proportion of ethnically minoritized populations is associated with higher rates of COVID-19 infection and death.^20^ Caregivers noticed their care recipients experienced accelerated physical and cognitive deterioration. This impact is in line with experimental evidence showing the link between engagement in activities and the preservation of physical and cognitive functioning in individuals with ADRD.^21,22^ This decline was also perceived to stem from a low access to cognitive medications during lockdown. Another factor that may have impacted physical and cognitive decline is the poor nutrition reported by some participants, which is a risk factor for many health conditions.^23^ Similar COVID-19-related nutrition findings have been shown among diverse populations around the world.^24^

Psychologically, chronic stress and depression were common among Latinos due to fear of infection, uncertainty, or isolation, and have been reported in other groups. ^13,25-27^ Research has shown a relationship between variables such as time of quarantine, constant fear of the care recipient’s death, or uncertainty and mental health outcomes.^25,28,29^ These impacts might be particularly strong among Latinos given their higher levels of socioeconomically-driven chronic stress.^30^ At the social level, the impact of the COVID-19 on the working situation among Latino caregivers has affected their finances when interrupted, or exposed them to infection risk and potentially stress as a consequence. Latinos may have been especially impacted by the work consequences of the pandemic, as they are overrepresented in the frontline workforce, tend to live in multigenerational homes, and have among the lowest average incomes.^31-33^

Social support was vital for most Latino families with ADRD during the pandemic. Many families moved in with their care recipient and took up more caregiving activities. However, some Latino families did not get family support during the pandemic and in fact, the pandemic reduced family visits, and worsened family dynamics in some cases. To make up for the lack of family support, some families capitalized on neighbors and friends. The family support many Latinos received in our study is in line with Latino’s family-centeredness.^34^ However, it is worth noting that in a recent national survey, Latino caregivers reported fewer family supports than non-Latino White and Black caregivers.^35^ Formal supports were also important among Latino families. Some Latinos also benefited largely from home care professionals, residences and all-inclusive care programs for the elderly which provided respite. However, the COVID-19 pandemic impacted the access to many formal sources of support.

Primary care was key in providing vaccines to many Latinos with ADRD and their families, reducing fear and increasing social support opportunities. However, COVID-19 signs confounded healthcare provider’s identification of ADRD signs and vice versa, which affected the quality-of-care Latinos with ADRD received. Flexible communication with informal and formal supports helped Latino families gain access to these while reducing the risk of COVID-19 infection. However, similar to other studies, the need to rely on remote communication intensified the digital divide.^13,26,27^ This divide may have especially affected Latino families with ADRD due to gaps in device ownership, connection to the internet, skills, and abilities, which may have been worsened by cognitive impairment, language barriers, and doubts about trustworthiness of the technology.

This study has some limitations. Remote recruitment and interviews increased the representation of participants in rural areas and other states. However, videocalls and phone calls led to some communication issues, which in some cases reduced the amount of information we could collect and affected the quality of the audio. The inability to conduct in-person recruitment and interviews may have excluded the most underserved individuals, who could have been contacted via health fairs before the pandemic started. We did not interview individuals with ADRD, which did not allow a full triangulation between them, their caregivers, and PCPs. While Latino caregivers tend to be women,^36^ these were over-represented in our study, likely also due to women’s higher likelihood to participate in health-related research.^37,38^ The sample size was relatively small and not probabilistic, which reduces the generalizability of the findings. As with most studies, individuals who participated in the study were motivated to participate. We do not know how much their discourse compares to those who decided not to participate.

This study has implications for public health. Given the efficacy of existing COVID-19 vaccines,^39^ ensuring access to ongoing boosters among Latinos with ADRD and their families will be needed. To do so, it will continue to be necessary to hold events at flexible times and days, convenient venues, and improve the communication with them by using a wide range of communication modalities (e.g., calling, texting, patient portals) in a linguistically and culturally appropriate way. The common stress and depression related to fear of COVID-19 infection, uncertainty, and confinement among Latino families with ADRD highlight the need improve access to mental health services in general and specifically during pandemics. An example of a potentially inexpensive intervention is layperson-delivered, empathy-oriented telephone call programs, which have shown to reduce loneliness, depression, and anxiety.^40^ These services can be provided by governments or charity organizations, and offered via primary care clinics, health departments and entities that identify those who are potentially in need. Other useful services may include cognitive and physical engagement activities to reduce confinement-related deterioration. Since stresses are cumulative and Latinos tend to have higher levels of socioeconomically-driven chronic stress,^30^ building a stronger and more accessible social and healthcare welfare systems will also be key. Our data suggests that decongestion of the mailing system and culturally-appropriate home-delivered foods may help reduce the COVID-19 pandemic’s impact on nutrition. Our data also suggests that promoting equity in gender roles may reduce the impact of future pandemics on nutrition among families where the caregivers are men. The impact of the pandemic on physical and cognitive decline and social supports and the crucial importance of families in the care of Latinos with ADRD highlight the need to provide them with financial support for their services, as well as respite when possible, and accessible caregiving support training. Home-care services have been essential in caring for some Latinos with ADRD and providing respite to their caregivers. However, these professions tend to be poorly paid, and hard to access. Home-care workers’ salary should be adjusted to the value of their service, and ideally covered by health insurance companies or government programs.

This work also has implications for future research. A PCP reported COVID-related death from some of his Latino patients during their yearly visit to Mexico. These visits are referred to as circular migration, in which large numbers of Mexicans head to the US in the spring and head to Mexico in the winter.^41^ Future research should explore the impact of COVID-19 among Latinos who do this type of migration. The fact that some PCPs suspected an unknown longer-term impact of the COVID-19 pandemic warrants further longitudinal research into this topic. Caregivers’ reports on healthcare providers’ confusion between ADRD and COVID-19 infection symptoms warrants research to improve diagnosis and severity assessments of both conditions.

## Conclusion

In this study, we have found that the COVID-19 pandemic has impacted Latino families with ADRD beyond infection and death, and informal and formal support resources have been crucial for their survival and quality of life. This pandemic has revealed many of the barriers that Latino families with ADRD face, and in most cases, this has exacerbated previous barriers.

However, with every crisis comes an opportunity for improvement, which will hopefully translate into improved conditions among Latino families with ADRD. These improved conditions might include more equitable access to health care and community services, a better quality of these services, subsidized formal and informal supports, and flexible hybrid means of communication.

## Data Availability

All data produced in the present study are available upon reasonable request to the authors

## Acknowledgements

The authors are thankful to Ms. Valois, RAIN, Children’s Mercy and other people and organizations that assisted with the recruitment of participants. The authors are also grateful to all the participants from this study, especially given the difficult circumstances due to the COVID-19 pandemic.

## References

1. Alzheimer’s Association. 2021 Alzheimer’s disease facts and figures. Alzheimer’s & Dementia. 2021;17(3)

2. Lin PJ, Emerson J, Faul JD, et al. Racial and Ethnic Differences in Knowledge About One’s Dementia Status. Journal of the American Geriatrics Society. 2020;68(8):1763–1770.

3. Barker WW, Luis C, Harwood D, et al. The effect of a memory screening program on the early diagnosis of Alzheimer disease. Alzheimer Disease & Associated Disorders. 2005;19(1):1–7.

4. Mehta KM, Yin M, Resendez C, Yaffe K. Ethnic differences in acetylcholinesterase inhibitor use for Alzheimer disease. Neurology. 2005;65(1):159–162.

5. Scharlach AE, Giunta N, Chow JC-C, Lehning A. Racial and ethnic variations in caregiver service use. Journal of Aging and Health. 2008;20(3):326–346.

6. Faison WE, Schultz SK, Aerssens J, et al. Potential ethnic modifiers in the assessment and treatment of Alzheimer’s disease: challenges for the future. International psychogeriatrics. 2007;19(3):539–558.

7. National Alzheimer’s Coordinating Center. National Alzheimer’s Coordinating Center: Query system. Accessed 12/21/2018, https://www.alz.washington.edu/WEB/naccquery.html

8. Connell CM, Roberts JS, McLaughlin SJ. Public opinion about Alzheimer disease among blacks, hispanics, and whites: results from a national survey. Alzheimer Disease & Associated Disorders. 2007;21(3):232–240.

9. Centers for Disease Control and Prevention. Demographic trends of COVID-19 cases and deaths in the US reported to CDC. 2022.

10. Painter EM, Ussery EN, Patel A, et al. Demographic characteristics of persons vaccinated during the first month of the COVID-19 vaccination program—United States, December 14, 2020–January 14, 2021. Morbidity and mortality weekly report. 2021;70(5):174.

11. Salgado de Snyder VN, McDaniel M, Padilla AM, Parra-Medina D. Impact of COVID-19 on Latinos: A Social Determinants of Health Model and Scoping Review of the Literature. Hispanic Journal of Behavioral Sciences. 2021;43(3):174–203.

12. Lines LM, Wiener JM. Racial and ethnic disparities in Alzheimer’s disease: A literature review. US Department of Health and Human Services, Assistant Secretary for Planning and Evaluation, Office of Disability, Aging and Long-Term Care Policy; 014.

13. Portacolone E, Chodos A, Halpern J, et al. The effects of the COVID-19 pandemic on the lived experience of diverse older adults living alone with cognitive impairment. The Gerontologist. 2021;61(2):251–261.

14. Schwandt TA. Dictionary of qualitative inquiry. Dictionary of qualitative inquiry. 2001:xxxiv, 281-xxxiv, 281.

15. Pollock T. The Difference Between Structured, Unstructured & Semi-Structured Interviews. Pridobljeno s https://www.oliverparkscom/blog-news/the-differencebetween-structured-unstructured-amp-semi-structured-interviews. 2019;

16. Basch CE. Focus group interview: an underutilized research technique for improving theory and practice in health education. Health Educ Q. Winter 1987;14(4):411–48.

17. Miles M, Huberman AM. Qualitative Data Analysis: A Source Book of New Methods. 1 ed. Sage Books; 1984:352.

18. Neergaard MA, Olesen F, Andersen RS, Sondergaard J. Qualitative description–the poor cousin of health research? BMC medical research methodology. 2009;9(1):1–5.

19. Taylor S, Treacy A. Just Dedoose it! Making mixed methods data analysis manageable. 2013:

20. Gorges RJ, Konetzka RT. Factors associated with racial differences in deaths among nursing home residents with COVID-19 infection in the US. JAMA network open. 2021;4(2):e2037431–e2037431.

21. Pitkälä K, Savikko N, Poysti M, Strandberg T, Laakkonen M-L. Efficacy of physical exercise intervention on mobility and physical functioning in older people with dementia: a systematic review. Experimental Gerontology. 2013;48(1):85–93.

22. Woods B, Aguirre E, Spector AE, Orrell M. Cognitive stimulation to improve cognitive functioning in people with dementia. Cochrane Database of Systematic Reviews. 2012;(2)

23. Sanders C, Behrens S, Schwartz S, et al. Nutritional status is associated with faster cognitive decline and worse functional impairment in the progression of dementia: the cache county dementia progression study 1. Journal of Alzheimer’s disease. 2016;52(1):33–42.

24. Penna PM, e Oliveira NMC, Castro LCV, Hermsdorff HHM. Consequences of Quarantine During the COVID-19 Pandemic on Food Intake and Body Weight: A Systematic Review. 2021;

25. Hawryluck L, Gold WL, Robinson S, Pogorski S, Galea S, Styra R. SARS control and psychological effects of quarantine, Toronto, Canada. Emerging infectious diseases. 2004;10(7):1206.

26. Cipolletta S, Morandini B, Tomaino SCM. Caring for a person with dementia during the COVID-19 pandemic: a qualitative study with family care-givers. Ageing & Society. 2021:1–21.

27. Vaitheswaran S, Lakshminarayanan M, Ramanujam V, Sargunan S, Venkatesan S. Experiences and needs of caregivers of persons with dementia in India during the COVID-19 pandemic—A qualitative study. The American journal of geriatric psychiatry. 2020;28(11):1185–1194.

28. Liew TM, Tai BC, Yap P, Koh GC-H. Comparing the effects of grief and burden on caregiver depression in dementia caregiving: a longitudinal path analysis over 2.5 years. Journal of the American Medical Directors Association. 2019;20(8):977-983. e4.

29. Groth N, Schnyder N, Kaess M, et al. Coping as a mediator between locus of control, competence beliefs, and mental health: A systematic review and structural equation modelling meta-analysis. Behaviour research and therapy. 2019;121:103442.

30. Brown LL, Mitchell UA, Ailshire JA. Disentangling the stress process: race/ethnic differences in the exposure and appraisal of chronic stressors among older adults. The Journals of Gerontology: Series B. 2020;75(3):650–660.

31. Goldman N, Pebley AR, Lee K, Andrasfay T, Pratt B. Racial and ethnic differentials in COVID-19-related job exposures by occupational standing in the US. PloS one. 2021;16(9):e0256085.

32. Semega J, Kollar M, Shrider EA, Creamer JF. Income and poverty in the United States: 2019. 2020. Current population reports. https://www.census.gov/content/dam/Census/library/publications/2020/demo/p60-270.pdf

33. Cohn DV, Passel JS. A record 64 million Americans live in multigenerational households. 2018.

34. Gallagher-Thompson D, Solano N, Coon D, Arean P. Recruitment and retention of Latino dementia family caregivers in intervention research: Issues to face, lessons to learn. The Gerontologist. 2003;43(1):45–51.

35. Rote SM, Angel JL, Moon H, Markides K. Caregiving across diverse populations: new evidence from the National Study of Caregiving and Hispanic EPESE. Innovation in aging. 2019;3(2):igz033.

36. National Alliance for Caregiving. Evercare® Study of Hispanic Caregiving in the U.S. 2008. https://www.unitedhealthgroup.com/content/dam/UHG/PDF/2008/Evercare_HispanicStudyFactSheet.pdf

37. Perales-Puchalt J, Shaw A, McGee JL, et al. Preliminary efficacy of a recruitment educational strategy on alzheimer’s disease knowledge, research participation attitudes, and enrollment among hispanics. Hispanic Health Care International. 2020;18(3):144–149.

38. Perales-Puchalt J, Barton K, Ptomey L, et al. Effectiveness of “Reducing Disability in Alzheimer’s Disease” among dyads with moderate dementia. Journal of Applied Gerontology. 2021;40(10):1163–1171.

39. Lopez Bernal J, Andrews N, Gower C, et al. Effectiveness of Covid-19 vaccines against the B. 1.617. 2 (Delta) variant. N Engl J Med. 2021:585–594.

40. Kahlon MK, Aksan N, Aubrey R, et al. Effect of layperson-delivered, empathy-focused program of telephone calls on loneliness, depression, and anxiety among adults during the COVID-19 pandemic: A randomized clinical trial. JAMA psychiatry. 2021;

41. Passel J, Cohn DV, Gonzalez-Barrera A. Net migration from Mexico falls to zero-and perhaps less: VI: Characteristics of Mexican-born immigrants living in the US. 2012.

